# Electronic Health Record-Based Estimation of Kansas City Cardiomyopathy Questionnaire Scores in Heart Failure

**DOI:** 10.64898/2026.04.03.26350138

**Authors:** Youngwon Kim, Wilson Lau, Nikhil Patel, Katherine Kendrick, Amy Wu, Ted Feldman, Ryan Ahern, Anand Oka

## Abstract

**Background:** The Kansas City Cardiomyopathy Questionnaire (KCCQ) is a validated patient-reported outcome measure for heart failure. However, its clinical utility is limited by incomplete and inconsistent data collection. We aimed to develop and validate machine learning models to estimate KCCQ overall summary scores from electronic health record (EHR) data.

**Methods:** We assembled a retrospective cohort of 10,889 heart failure patients with recorded KCCQ scores from the Truveta database. Predictor features were derived from structured EHR variables across 13 historical time windows (15-360 days). Multiple regression algorithms were evaluated, followed by SHapley Additive exPlanations (SHAP)-based feature reduction and nested cross-validation for hyperparameter optimization. Model performance was assessed using the coefficient of determination (*R*^2^), mean absolute error (MAE), and ordinal discrimination and calibration for categorical severity classification.

**Results:** Histogram-based gradient boosting (HGB) with HGB-SHAP feature selection achieved the strongest performance, reducing feature dimensionality by more than 94% while maintaining estimation accuracy. The 240-day window performed best (*R*^2^=0.522, MAE=12.485). For categorical severity classification, the model demonstrated strong ordinal discrimination (mean ordinal AUROC=0.850). Quantile-based calibration improved classification balance, increasing the F1-score for the most severe category (KCCQ<25) from 0.180 to 0.428 and the quadratic weighted kappa from 0.601 to 0.640. Longer EHR observation windows were associated with improved prediction performance.

**Conclusion:** Machine learning models can estimate KCCQ scores from routine EHR data with clinically meaningful accuracy and strong discriminatory performance. This approach may help extend assessment of patient-reported health status to populations in which survey-based data are incompletely captured, supporting population-level cardiovascular outcomes assessment and risk stratification in heart failure care.

**What is Known:**

- Patient-reported outcomes, such as the Kansas City Cardiomyopathy Questionnaire (KCCQ), are essential for assessing health status in patients with heart failure but are inconsistently collected in routine clinical practice, limiting their use for population-level monitoring and outcomes assessment.
- Despite their clinical importance, prior studies have primarily used KCCQ scores as predictors of downstream clinical outcomes, and few approaches have been developed to estimate KCCQ scores directly from electronic health record (EHR) data.

**What the Study Adds:**

- This study presents a rigorously validated machine learning framework for estimating KCCQ scores from routinely collected EHR data, achieving clinically meaningful accuracy across multiple temporal windows with substantial feature reduction.
- The proposed approach incorporates post-hoc calibration to improve identification of patients with severe functional impairment, supporting scalable assessment of patient-reported health status in settings where survey data are incomplete or unavailable.

## 1. Introduction

Cardiovascular diseases (CVD) remain a major contributor to death in the United States, with heart disease-related mortality rising in recent years [1]. In 2022, heart failure, specifically, was cited on 457,212 death certificates, accounting for 13.9% of all U.S. deaths [2]. This trend indicates the critical need for improved approaches to risk assessment and health status measurement that capture the multidimensional impact of CVD and support population-level monitoring and clinical decision-making.

One widely used tool for assessing health status in heart failure is the Kansas City Cardiomyopathy Questionnaire (KCCQ), a patient-reported outcome (PRO) measure qualified by the U.S. Food and Drug Administration (FDA) as a clinical outcome assessment [3]. The KCCQ is a validated instrument designed to assess quality of life, symptom burden, and physical and social functioning in individuals with heart failure and valvular heart disease [4, 5, 6, 7]. These domains are vital for evaluating disease severity, stratifying patient populations in clinical trials, and guiding treatment decisions [8, 9].

KCCQ scores have well-established prognostic value. Overall summary scores are strongly associated with outcomes such as hospitalization and mortality across various stages of heart failure [7, 10]. For example, a 5- or 10-point change in the score represents a clinically meaningful shift in the risk for subsequent cardiovascular death or hospitalization [4, 11, 12]. Patients with KCCQ scores below 25 face approximately three times greater risk of death or hospitalization compared to those with similar clinical characteristics but scores above 75 [13, 14]. Beyond its prognostic utility, the KCCQ is often used as a health status endpoint in intervention studies, including trials of transcatheter tricuspid valve interventions (TTVI) [15, 16]. More broadly, KCCQ scores can function as signals for clinical deterioration, prompting proactive interventions such as medication adjustments, intensified monitoring, or rehabilitation referrals-potentially preventing hospitalizations and other adverse events [9, 11, 17, 18].

Despite its clinical and research importance, the KCCQ’s practical application is limited by its reliance on patient completion. Critically ill patients, those with competing demands, and busy coordinators often fail to provide complete data [19]. Additional barriers to completion include patient dissatisfaction, perceived burden, time constraints, language or literacy challenges, or even death prior to survey administration [20, 21, 22]. As a result, KCCQ data are frequently sparse not only within electronic health records (EHRs), but also in clinical registries [23, 21, 24]. This sparsity can hinder comprehensive patient assessment, introduce bias into research findings, and limit accurate risk stratification for a substantial portion of the heart failure population.

Routinely collected EHR data offer a potential approach to address these limitations. EHRs contain rich, longitudinal clinical information on symptoms, comorbidities, laboratory results, and healthcare utilization that reflect many of the constructs captured by the KCCQ. Estimating KCCQ scores from these data could extend assessment of patient-reported health status to individuals who do not complete questionnaires, thereby supporting more inclusive population-level monitoring of heart failure outcomes.

Prior studies applying machine learning (ML) in heart failure have typically used KCCQ scores as predictors of other outcomes (e.g., mortality, hospitalization) or have relied on limited sets of manually selected clinical variables. [14, 25, 26]. In contrast, estimating KCCQ scores directly from high-dimentional, longitudinal EHR data represents a distinct and less explored problem. Addressing this gap may support broader, scalable assessment of health status beyond settings in which surveys are consistently administered.

In this study, we develop and validate ML models to estimate KCCQ overall summary scores using real-world EHR data from a large and diverse cohort of patients with heart failure. We evaluate estimation accuracy, categorical severity classification, and alignment with clinically meaningful thresholds to assess whether EHR-based approaches can support scalable and clinically relevant measurement of heart failure health status. This framework has the potential to enhance population-level cardiovascular outcomes assessment and inform future applications of extending PRO measurement to settings in which direct survey data are incomplete or unavailable.

## 2. Objectives

We aim to develop and validate ML models that predict the KCCQ overall summary score from historical, de-identified EHR data. Using a retrospective cohort from the Truveta database, we evaluate both predictive accuracy and clinical relevance in patients with heart failure.

**RQ1.** How accurately can ML models estimate KCCQ overall summary scores using available patient data from EHRs?

**RQ2.** To what extent do predicted KCCQ scores align with clinically meaningful categorical classifications of heart-failure severity?

**RQ3.** Can EHR-based KCCQ estimates identify patients with poor health status, and are the features driving these estimates consistent with known risk factors for low KCCQ scores?

## 3. Methods

### Data source and study cohort

De-identified patient data were obtained from the Truveta database, a comprehensive repository aggregating EHR data across multiple U.S. health systems. The database represents a large and diverse patient population-over 120 million individuals-and includes care provided in more than 900 hospitals and 20,000 clinics across the U.S. [27]. We identified patients with heart failure who had at least one recorded KCCQ overall summary score between 2012 and 2025 (Figure 1). KCCQ scores were extracted using validated concept codes (SNOMED CT: 471011000124100; LOINC: 86924-8 and 71940-1; Supplemental Methods S1). Data sharing and code availability are described below.

**Figure 1:**
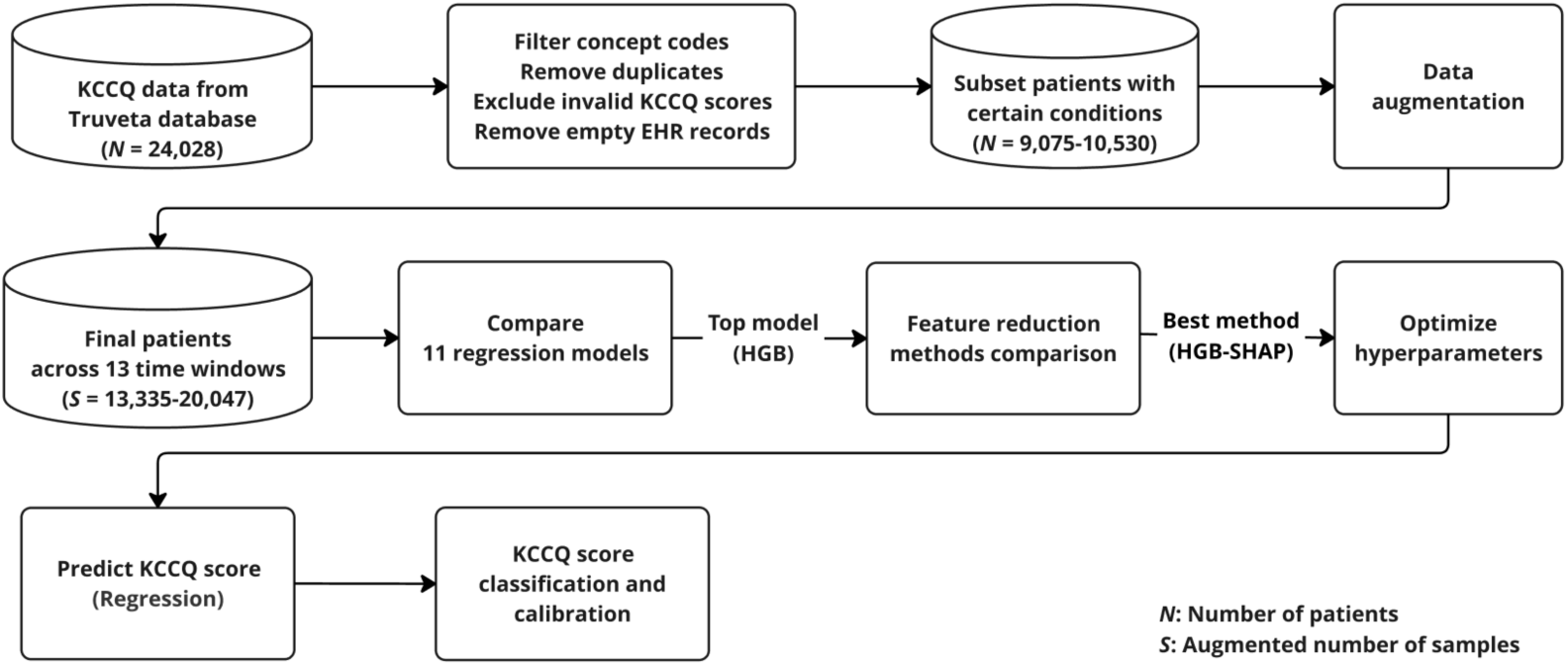
KCCQ study overview.

We excluded: (1) records with missing KCCQ scores, (2) KCCQ scores outside the clinically valid range (0-100), (3) duplicate entries (defined as multiple records for the same patient on the same date), and (4) patients lacking any prior EHR history (observations, conditions, lab results) preceding the KCCQ assessment date. After applying these criteria, the final analytic cohort comprised 10,889 unique patients with at least one valid KCCQ overall summary score and associated historical EHR data. The final cohort had mean KCCQ scores of 54.04-55.86, a mean age of 74-77 years, and was 40-41% female; detailed characteristics are reported in Supplemental Methods S2.

### Outcome definition and severity categories

The primary outcome was the KCCQ overall summary score, which ranges from 0 to 100 with higher values indicating better health status over a two-week recall period. Item-level responses were largely unavailable, and the specific KCCQ version (KCCQ-12 vs KCCQ-23) was not consistently specified for all records. Given prior evidence demonstrating strong correlation between overall summary scores from the two versions, all overall scores were treated as a common endpoint [8, 9]. For clinical interpretation, KCCQ scores were additionally categorized into established severity strata: very poor to poor (0-25), poor to fair (25-50), fair to good (50-75), and good to excellent (75-100) [9]. For additional details on the KCCQ instrument, version handling, and domain structure, see Supplemental Methods S3.

### Data windows and predictor features

To assess the influence of longitudinal EHR context on estimation performance, predictor variables were constructed from patients EHR histories across 13 time windows preceding each KCCQ assessment (15 to 360 days). Patients could contribute multiple KCCQ assessments, with each assessment treated as a distinct clinical snapshot paired with features derived from its corresponding lookback window (Supplemental Methods S4). Accordingly, model development and evaluation were conducted at the assessment level rather than the patient level.

Predictor features were derived from structured EHR data within each window, including demographics (one-hot encoded), clinical conditions (frequency counts), and laboratory and vital sign measurements (window-specific summaries). Feature construction followed a standardized, data-driven pipeline designed for high-dimensional and sparse EHR inputs (Supplemental Methods S5).

### Model development and validation

We developed and evaluated regression models to estimate continuous KCCQ scores using a three-stage workflow: (1) initial screening of candidate regression model families, (2) comparison of feature reduction strategies, and (3) hyperparameter optimization of the selected model. Model development and evaluation were conducted within a nested cross-validation (CV) framework to prevent information leakage, with performance summarized using the coefficient of determination (*R*^2^) and mean absolute error (MAE).

In the model screening stage, histogram-based gradient boosting (HGB) consistently outperformed alternative regression approaches across time windows and was selected as the base model. We then compared multiple feature reduction strategies, including SHapley Additive exPlanations (SHAP)-based rankings, model-derived feature importances, and penalization-based methods (Lasso and Elastic Net). Among these approaches, HGB-derived SHAP feature ranking demonstrated stable performance across CV folds, reducing the feature space to under 5.5% of the original while maintaining performance comparable to models trained on the full predictor set. Accordingly, the final model consisted of an HGB regressor with HGB-based SHAP feature selection and hyperparameters optimized through nested cross-validation. Full methodological details are provided in Supplemental Methods S6.

### Clinical stratification and calibration

For clinical applicability, continuous KCCQ estimates were converted into ordinal severity categories using standard clinical thresholds. Because naïve post-hoc binning of regression outputs can lead to mismatches between predicted and observed category prevalence, we evaluated multiple calibration approaches and selected a quantile-binning approach. In this procedure, category thresholds were derived from empirical quartiles of the training outcome distribution and applied to corresponding test predictions, ensuring calibration without information leakage (Supplemental Methods S7). Categorical performance was evaluated using accuracy, precision, recall, F1-score, ordinal area under the receiver operating characteristic curve (AUROC), and quadratic weighted kappa (QWK).

## 4. Results

### Final model performance with optimized HGB

Model performance is reported at the assessment level, with each KCCQ measurement treated as a distinct patient-timepoint sample derived from its corresponding time window. We proceeded with HG) using HGB-SHAP feature selection for final model development. After hyperparameter optimization, HGB with HGB-SHAP consistently reduced the original 11,833-19,180 features to 648-796 features (3.6-5.5% of the initial sets; Table 1). This substantial feature reduction maintained predictive performance while improving computational efficiency.

**Table 1:**
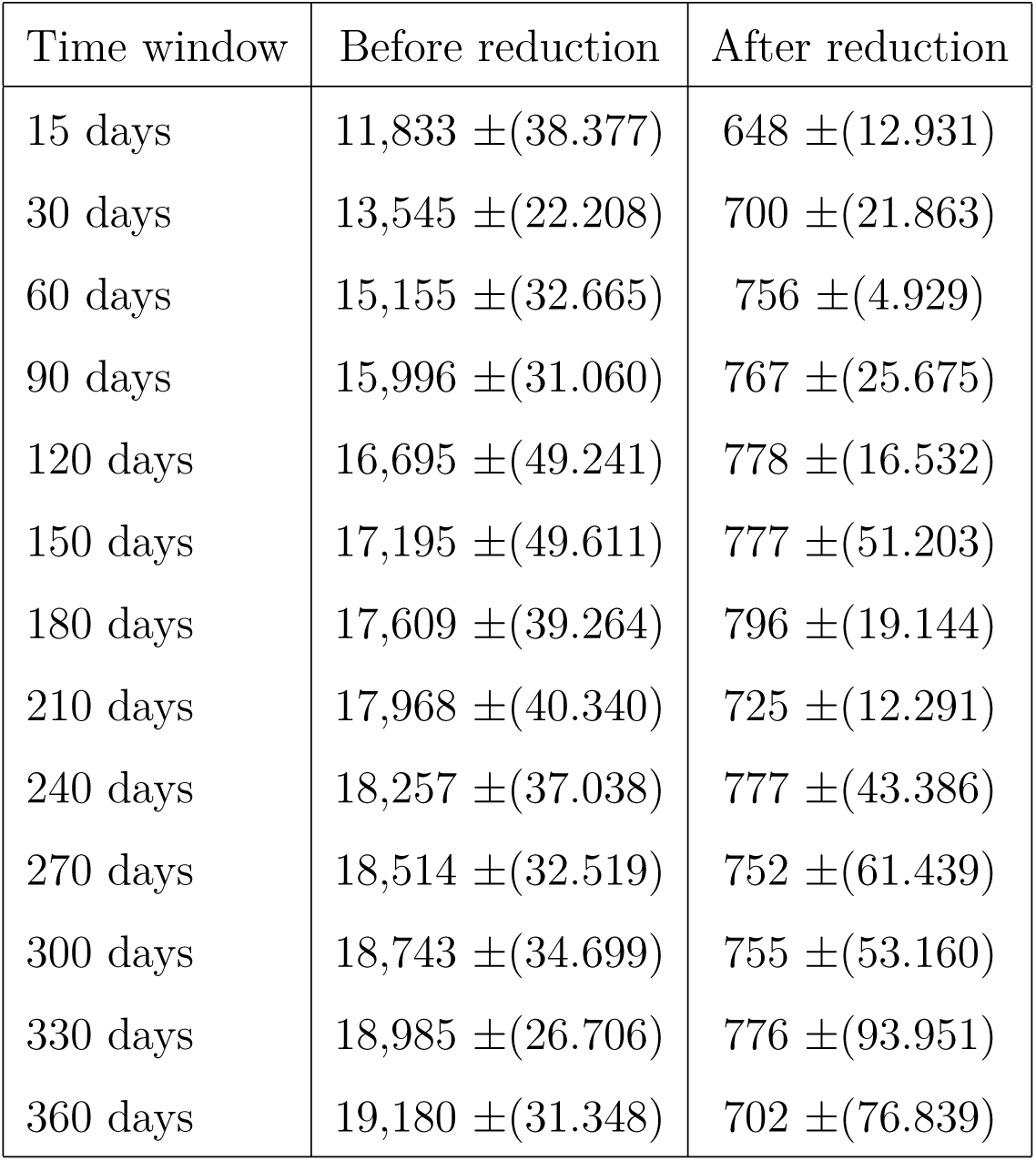
Feature count before and after reduction across 13 time windows (mean *±* SD across outer cross-validation folds)

| Time window | Before reduction | After reduction |
| --- | --- | --- |
| 15 days | 11,833 ±(38.377) | 648 ±(12.931) |
| 30 days | 13,545 ±(22.208) | 700 ±(21.863) |
| 60 days | 15,155 ±(32.665) | 756 ±(4.929) |
| 90 days | 15,996 ±(31.060) | 767 ±(25.675) |
| 120 days | 16,695 ±(49.241) | 778 ±(16.532) |
| 150 days | 17,195 ±(49.611) | 777 ±(51.203) |
| 180 days | 17,609 ±(39.264) | 796 ±(19.144) |
| 210 days | 17,968 ±(40.340) | 725 ±(12.291) |
| 240 days | 18,257 ±(37.038) | 777 ±(43.386) |
| 270 days | 18,514 ±(32.519) | 752 ±(61.439) |
| 300 days | 18,743 ±(34.699) | 755 ±(53.160) |
| 330 days | 18,985 ±(26.706) | 776 ±(93.951) |
| 360 days | 19,180 ±(31.348) | 702 ±(76.839) |

Average predictive performance across the 13 time windows is shown in Figure 2. Compared with unoptimized HGB, hyperparameter tuning improved overall *R*^2^ by 0.002-0.016 and reduced MAE by 0.085-0.223 consistently across all time windows.

**Figure 2:**
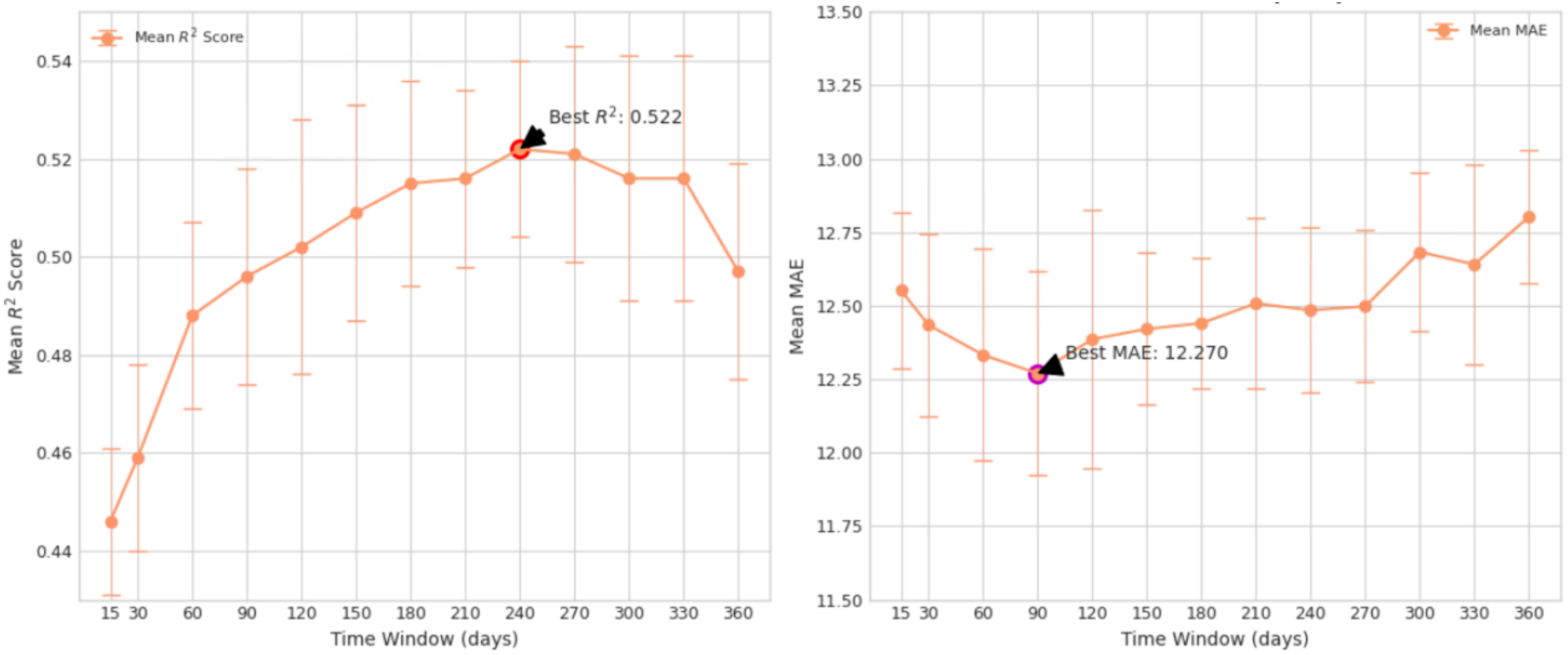
Overall performance after HGB–SHAP feature selection and evaluation with nested cross–validation, supporting generalizability.

Performance varied by observation window length. The 15-day window yielded the weakest performance, despite aligning with the KCCQs two-week recall period. The 240-day window achieved the highest explained variance (*R*^2^ = 0.522) with an MAE of 12.485, explaining 52.2% of score variability. The lowest MAE (12.270) was observed at 90 days, although its *R*^2^ (0.496) was slightly lower. Performance stabilized across longer windows (60-360 days), with *R*^2^ values ranging from 0.488 to 0.522 and MAE values from 12.270 to 12.803, with no statistically significant differences relative to the best-performing window (240 days; *p >* 0.05 based on paired Wilcoxon signed-rank tests across outer CV folds).

### Clinical applicability: Categorical severity prediction

To assess clinical applicability, we converted continuous KCCQ predictions from the best-performing 240-day model into clinically defined severity categories: Very Poor to Poor (0-25), Poor to Fair (25-50), Fair to Good (50-75), and Good to Excellent (75-100). Model performance was evaluated on 14,260 samples across five held-out test sets from nested CV.

As shown in Table 2, the initial categorical predictions demonstrated strong ordinal discrimination (mean ordinal AUROC = 0.850) but limited sensitivity for the most severe category, with a low F1-score for the Very Poor to Poor group (F1 = 0.180). Given the clinical importance of identifying patients with severe functional impairment, we applied post-hoc calibration to rebalance category assignments with preserving the underlying continuous predictions.

**Table 2:**
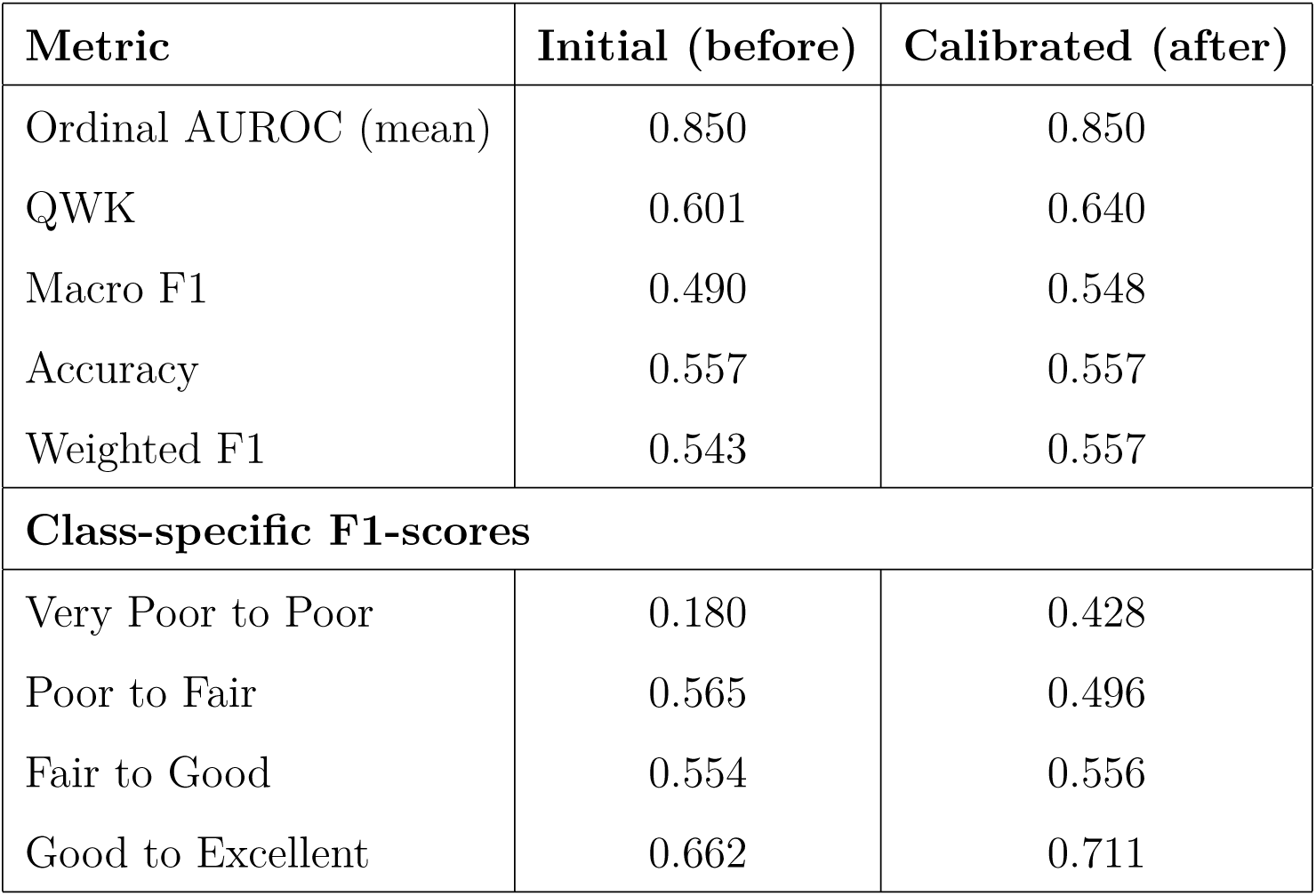
Performance summary at 240 days before and after calibration.

Following calibration, class-balanced and ordinal agreement metrics improved. Macro F1, which weights all severity categories equally, increased from 0.490 to 0.548, while QWK, reflecting agreement across ordered categories, improved from 0.601 to 0.640. In contrast, overall accuracy and weighted F1both influenced by class prevalenceremained largely unchanged after calibration (changes of +0.001 and +0.015, respectively). Examination of class-specific F1-scores (Table 2) shows the intended effect of calibration: the F1-score for the Very Poor to Poor category increased from 0.180 to 0.428, and performance for the Good to Excellent category improved from 0.662 to 0.711. These gains were accompanied by modest changes in the intermediate categories (0.069 for Poor to Fair and 0.002 for Fair to Good).

Figure 3 provides complementary views of model performance. Because calibration adjusted only categorical thresholds, ROC and PR curves based on continuous predicted KCCQ values remained unchanged. ROC curves (Figure 3 A) showed good discrimination for identifying severe impairment at the primary screening threshold (KCCQ < 25; AUC = 0.822), comparable performance at KCCQ < 50 (AUC = 0.817), and higher discrimination at KCCQ < 75 (AUC = 0.911). PR curves (Figure 3 B) contextualized these results under observed class prevalences, with average precision increasing from 0.452 for severe impairment (KCCQ < 25; prevalence 10%) to 0.970 for KCCQ < 75 (prevalence 79%). Confusion matrices before and after calibration (Figure 3 C-D) illustrate improved capture of the most severe category while preserving the overall classification structure across KCCQ severity levels.

**Figure 3:**
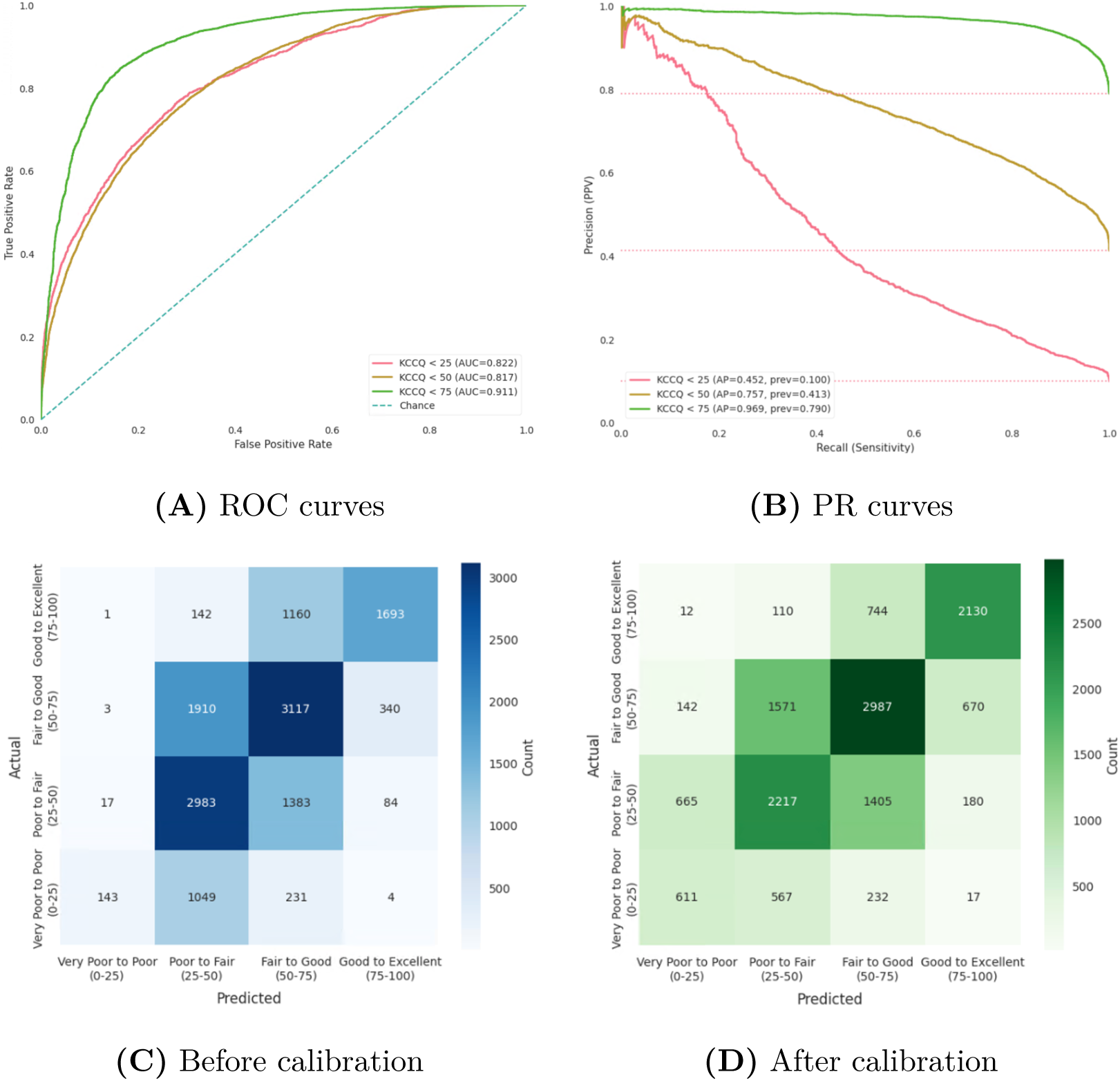
Overall performance of the 240-day model. (A) ROC curves for identifying clinically impaired patients using KCCQ thresholds (< 25, < 50, and < 75; (unchanged by threshold calibration); (B) PR curves for the same thresholds; (C–D) Confusion matrices for four-category KCCQ severity classification before and after quantile-based threshold calibration

### Key features influencing KCCQ Scores

#### Global insights from training set

We examined the top 25 features that most strongly influenced KCCQ predictions in the 240-day model by analyzing global SHAP-based importance scores (Figure 4). Our analysis showed that the model’s predictions were driven by a diverse set of features that can be grouped into distinct clinical domains.

**Figure 4:**
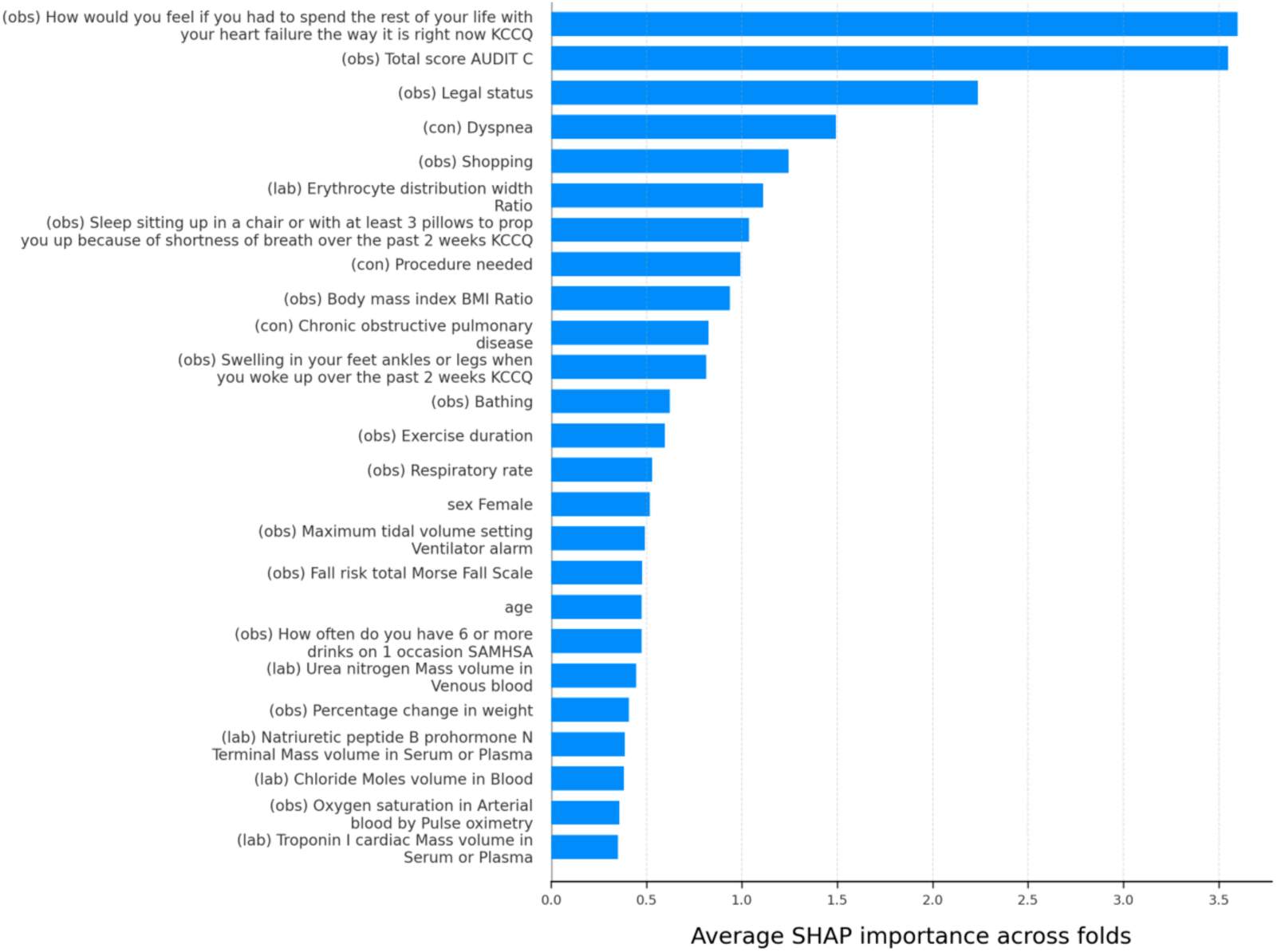
Global feature importance estimated by average SHAP values across all predictions (240 day)

Three individual KCCQ items emerged as influential predictors: "How would you feel if you had to spend the rest…," "Swelling in your feet, ankles, or legs…," and "Sleep sitting up in a chair or with at least…". These items directly capture symptom burden, reflecting the core dimensions of heart failure impact on daily life. Notably, these KCCQ item scores were highly sparse in our dataset, with approximately 80% of values missing.

Lifestyle indicators were also important predictors. Alcohol use variables such as the AUDIT-C total score and frequency of heavy drinking episodes and activities such as shopping, bathing, and exercise duration captured functional capacity and quality of life domains contributed to predictions, capturing functional capacity and quality-of-life domains closely aligned with the KCCQ framework.

Respiratory comorbidities-including chronic obstructive pulmonary disease (COPD), dyspnea, respiratory rate, and ventilator tidal volume alarm settings-were also strong contributors. Several physiological and laboratory measurements further influenced predictions, including erythrocyte distribution width ratio, blood urea nitrogen, natriuretic peptide levels, body mass index, oxygen saturation, chloride, and troponin I. Additionally, demographic variables such as legal status, sex (female), and age also contributed to KCCQ predictions, reflecting the influence of social and biological context on patient-reported health status.

#### Local insights from the test set (bottom 25%)

We next examined SHAP-based feature importance restricted to test set patients in the bottom 25% of predicted KCCQ scores, representing individuals with the poorest heart health status (Figure 5). Compared with the global feature importance results, the overall feature importance profile in this subgroup was largely consistent, with similar categories of patientreported outcomes, functional measures, comorbidities, laboratory values, and demographic variables contributing to predictions. However, differences were observed in the relative ordering of features. Alcohol use (AUDIT-C total score), functional limitations (e.g., shopping and bathing), and demographic variables (age and sex) ranked higher in importance in the bottom 25% subgroup, while individual KCCQ items showed slightly reduced relative dominance compared with the global analysis.

**Figure 5:**
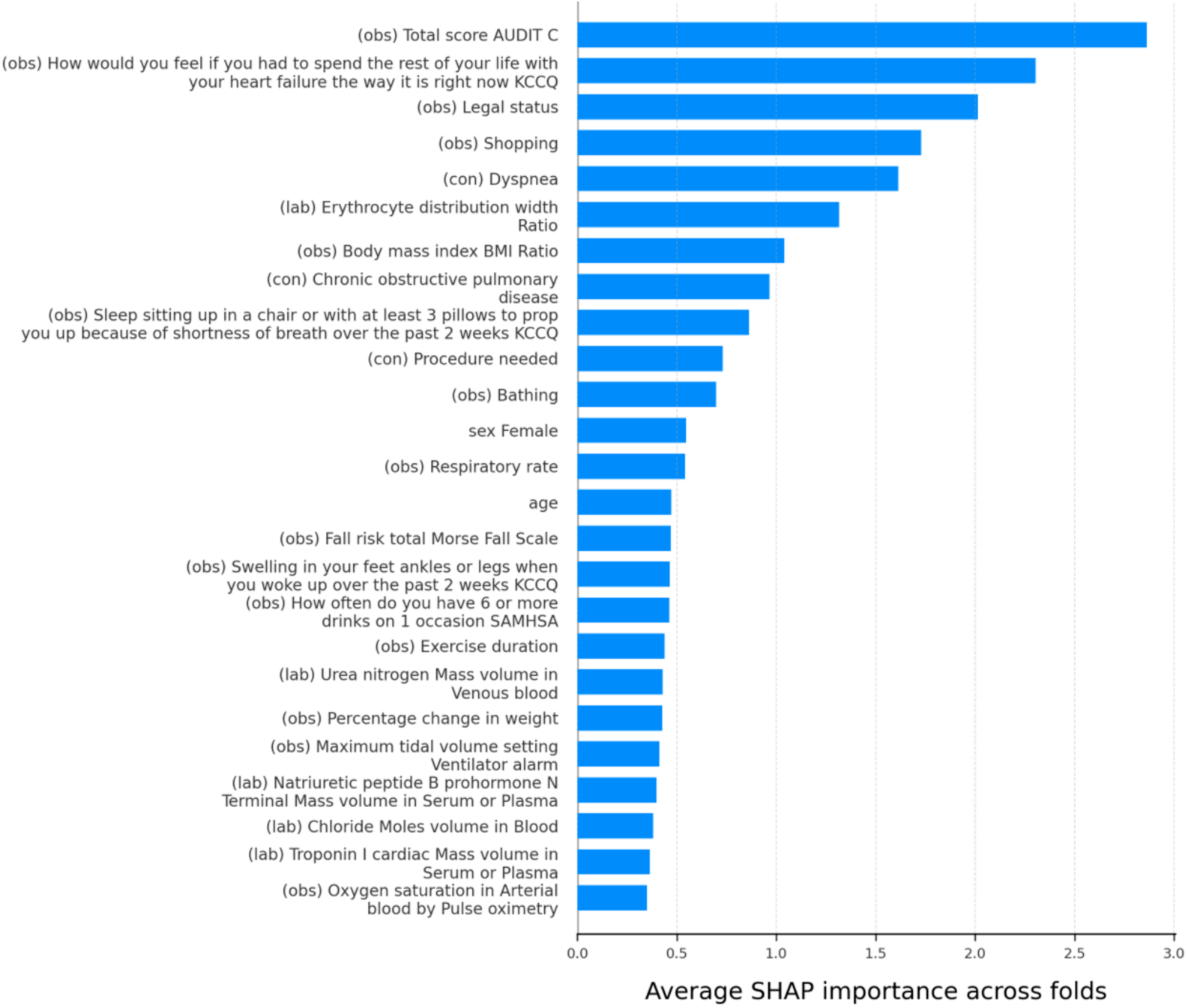
Local feature importance estimated by average SHAP values for test set patients in the bottom 25% of predicted KCCQ scores (240 day)

## Discussion

We developed and validated a machine learning framework for predicting KCCQ scores using EHR data from the Truveta database. To our knowledge, few studies have applied machine learning to infer KCCQ overall scores directly from real-world EHR data, and our work provides a comprehensive benchmark across time windows, feature reduction strategies, and post-hoc calibration. More broadly, these results indicate that general EHR datawithout reliance on disease-specific survey instrumentscan produce clinically meaningful estimates of patient-reported heart-health status.

Three main contributions emerged: (1) temporal analysis identified optimal observation windows, (2) calibration improved clinically relevant categorical severity classification, and (3) SHAP-based feature importance characterized the multidomain features associated with KCCQ predictions.

### Temporal windows and predictive performance

Our temporal analysis revealed important insights into the relationship between observation window length and predictive accuracy. Despite the KCCQs 2-week recall period [9], the 15-day window produced the weakest results across all models. In contrast, predictive performance improved with longer observation windows and peaked at 240 days (*R*^2^ = 0.522). This pattern suggests that effective prediction requires capturing broader longitudinal patterns rather than short-term symptom fluctuations, reflecting the cumulative nature of health experiences and treatment responses that shape KCCQ scores.

From a statistical perspective, these results represent strong predictive performance in a clinical context. In many real-world clinical prediction studies, a an explained variance (*R*^2^) of 15% is often considered meaningful in clinical studies given the inherent complexity of predicting patient outcomes [28, 29]. Our peak explained variance of 52% significantly exceeds this established benchmark and compares favorably to prior work. For example, a study mapping the MacNew-7D instrument against KCCQ achieved *R*^2^ values of 0.480-0.600 [30]. That our EHR-based approach-using longitudinal, retrospective real-world clinical data rather than heart disease-specific measures-achieved comparable performance demonstrates the clinical relevance of routinely collected EHRs.

However, the added benefit of the 240-day window over shorter periods was modest. Observation windows of 60 to 90 days provided nearly equivalent accuracy while demanding fewer computational resources and aligning more closely with typical clinical follow-up schedules. As such, these shorter windows may represent a practical compromise for real-world implementation, balancing predictive performance, data availability, and operational constraints, particularly in settings where many patients have limited hospital visit histories.

### Calibration for clinically actionable severity classification

The strong ordinal discrimination observed prior to calibration (mean ordinal AUROC = 0.850) indicates that the model reliably captured relative differences in patient functional status across the severity spectrum [31]. This level of discrimination supports the use of the model for continuous estimation and ranking of KCCQ scores. However, when continuous predictions were mapped to discrete severity categories, categorical performance was uneven across outcome levels, with particularly limited identification of patients in the most severe category.

Post-hoc calibration was therefore applied not to improve discrimination, but to enhance the clinical usability of categorical predictions while preserving the underlying continuous outputs. By adjusting category thresholds without altering prediction rankings, calibration redistributed sensitivity toward clinically consequential extremes of disease severity. As a result, per-class F1-scores increased substantially for the Very Poor to Poor category (+0.248) and improved for the Good to Excellent category (+0.049), while only modest changes were observed in intermediate groups (0.069 and +0.003). These trade-offs reflect a clinically meaningful rebalancing, prioritizing improved recognition of the most vulnerable patients over marginal losses in less critical categories.

From a clinical perspective, this distinction is important. Accurate ordinal ranking supports longitudinal monitoring and risk stratification, whereas categorical classification is often required for decision-making, triage, and population-level screening. The calibration strategy adopted here reflects a deliberate prioritization of clinical relevance over marginal gains in aggregate accuracy, acknowledging that misclassification of severely impaired patients carries greater potential consequences than uncertainty within intermediate severity levels.

At the aggregate level, calibration improved class-balanced performance, as reflected by increases in macro F1 (+0.053) and balanced recall (+0.066), while overall accuracy and weighted F1 remained largely unchanged. This pattern underscores the importance of macro-averaged metrics for evaluating clinically oriented models, as prevalence-weighted measures tend to obscure performance gains in minority but high-risk groups. The accompanying increase in QWK (to 0.640) further indicates improved agreement across ordered severity categories, reaching the threshold for substantial agreement [32].

Taken together, these findings indicate that calibration serves as a pragmatic, clinically motivated refinement rather than a corrective adjustment. By aligning categorical outputs with clinical priorities while preserving strong discriminative performance, post-hoc calibration enhances the translational value of EHR-based patient-reported outcome models, particularly in settings where identifying patients at highest risk is a primary objective.

### Insights from feature importance

The feature importance analysis indicated that the model utilized a broad range of information available in existing EHRs – including symptom-related variables, functional measures, objective biomarkers, and demographic characteristics – to generate KCCQ predictions and how these patterns may inform clinical applicability.

The presence of individual KCCQ items among the top predictors confirms that item-level responses, when available, strongly inform the composite score [8]. While using individual KCCQ item scores as features to predict the KCCQ total score could potentially raise the theoretical concern of information leakage, this risk is mitigated in the present setting by the high degree of sparsity in these variables (approximately 80% missing). In practice, this sparsity underscores a key strength of our approach: the model’s ability to generate predictions primarily from routinely collected EHR data rather than relying on patient-reported questionnaire responses.

Beyond KCCQ items, influential features spanned multiple clinical domains. Lifestyle and functional indicators (e.g., AUDIT-C alcohol use, daily activity measures) suggest that routinely collected EHR data can approximate domains traditionally assessed through PROs. Similarly, the contribution of respiratory comorbidities and physiological markers reflects the multidomain burden of heart failure, consistent with prior evidence that heart health status is shaped by both clinical and functional dimensions [33]. Demographic contributions, such as sex, age, legal status, further emphasize the role of social and biological context on health-related quality of life [34].

Comparisons between the global feature importance results and the bottom 25% subgroup revealed largely overlapping sets of influential features, with differences primarily in relative ranking rather than feature composition. This consistency suggests that the model relied on similar types of information across the overall population and among patients with poorer heart health, while placing greater relative emphasis on functional and lifestyle indicators in the lower KCCQ subgroup. In particular, heavy alcohol use showed increased importance among patients with worse predicted heart health [35]. As with all feature attribution methods, these findings reflect associations rather than causal relationships and should be interpreted accordingly.

Feature importance patterns may vary across time windows. The patterns observed at 240 days represent one snapshot of a dynamic relationship between EHR data and KCCQ scores. Nonetheless, the recurring presence of symptom, functional, respiratory, and laboratory domains suggests that EHR-based models may capture clinically relevant aspects of heart-related health status [36, 37] even when survey data are incomplete. These findings support the potential utility of EHR-based approaches to extend estimation of patient-reported health status to populations that are less frequently captured through traditional questionnaire-based assessments, though further validation is necessary in broader and more diverse cohorts.

### Relation to prior work and methodological considerations

Our findings contribute to ongoing discussions about the role of PROs in predictive modeling, demonstrating that routinely collected EHR data can be used to estimate KCCQ scores in settings where direct survey responses are incomplete or unavailable. Previous studies have underscored both the opportunities and the challenges of incorporating PROs into clinical prediction models [38, 39].

Within this context, tree-based models-especially HGB-showed strong performance, consistent with prior evidence that such approaches can capture complex, non-linear relationships in high-dimensional, sparse EHR data [40]. At the same time, their flexibility increases the risk of overfitting. The use of nested CV and post-hoc calibration reflects an emphasis on methodological rigor and clinical reliability, reducing the likelihood that observed performance was driven by idiosyncratic noise. Calibration further aligned categorical outputs with clinically meaningful severity distinctions, improving usability for patients with severe impairment while preserving discriminative performance.

Feature importance analysis also situates our work within existing research. Prior studies have emphasized the prognostic significance of multidomain factors in heart failure, including functional capacity, comorbid respiratory disease [41], and laboratory biomarkers such as EDR, natriuretic peptides, and troponin I [42, 43, 44]. Our findings are consistent with this literature but extend it by showing that models trained on general EHR datanot limited to disease-specific surveyscan cover clinically similar domains of relevance. These findings suggest that model trained on routinely collected EHRs may approximate constructs traditionally measured through PROs, supporting their complementary role in outcome modeling.

### Future directions

Future work should examine additional clinical variables that may moderate or mediate the relationship between PROs and objective endpoints, such as hospitalization or mortality. As the Truveta database continues to expand, repeating these analyses with larger and more diverse cohorts may further strengthen performance and generalizability.

An important consideration is that certain populations may have limited access to, or lower completion rates of, KCCQ surveys due to barriers such as language, literacy, digital access, or patterns of healthcare engagement. With appropriate external validation, EHR-derived KCCQ estimates may help extend health status assessment, longitudinal monitoring, and risk stratification to patients who do not routinely complete survey-based instruments. Such approaches could support earlier identification of patients at risk for functional decline while helping to address gaps in cardiovascular quality assessment. Future research should specifically examine model performance across demographic subgroups to assess whether this approach improves equity in health status measurement or risks perpetuating existing disparities.

More broadly, this framework may be applicable to other patient-reported health constructs, although such applications would require condition-specific development and validation.

### Limitations

This study relied on retrospective EHR data, which are subject to coding variability, missingness, and incomplete capture of patient histories, potentially limiting generalizability beyond health systems with similar data structures and populations. External validation in independent cohorts is needed to confirm robustness across diverse clinical settings.

In addition, only overall KCCQ summary scores were available, without consistent item-level responses or differentiation between KCCQ versions, which may limit predictive precision. Post-hoc calibration improved identification of severe impairment but involved trade-offs in intermediate categories, and models were restricted to structured EHR data, excluding potentially informative sources such as social determinants of health or unstructured clinical data.

## Nonstandard Abbreviations and Acronyms

KCCQ: Kansas city cardiomyopathy questionnaire
CVD: Cardiovascular diseases
FDA: Food and Drug Administration
ML: machine learning
EHR: electronic health record
SHAP: SHapley Additive exPlanations
MAE: mean absolute error
PRO: patient-reported outcome
TTVI: transcatheter tricuspid valve interventions
SNOMED CT: systematized nomenclature of medicine-clinical terms
LOINC: logical observation identifiers names and codes
CV: cross validation
QWK: quadratic weighted kappa
AUROC: area under the receiver operating characteristic
PR: precision-recall
RF: random forest
KNN: *k*-nearest neighbors regression
SVR: support vector regression
AdaBoost: adaptive boosting
HGB: histogram-based gradient boosting
XGBoost: extreme gradient boosting
LightGBM: light gradient-boosting machine regression
COPD: chronic obstructive pulmonary disease
EDR: Erythrocyte distribution width ratio

## Acknowledgments

We gratefully acknowledge the contributions and support from Md Enamul Haque and Jay Nandury.

## Sources of Funding

This work was funded by Truveta Inc.

## Disclosures

Y. Kim, M. E. Haque, J. Nandury, and other listed Truveta-affiliated authors are employees of Truveta Inc. T. Feldman is an employee of Edwards Lifesciences. The authors report no other disclosures.

## Declaration of Competing Interest

The authors are employees of Truveta Inc., and this work was conducted as part of their research activities at the company. T. Feldman is an employee of Edwards Lifesciences. The authors declare no additional competing financial interests or personal relationships that could have influenced the work reported in this paper.

## Declaration of generative AI use

During the preparation of this work, large language models were used to assist with code refinement for data preparation and analysis, improve data visualizations, and enhance the clarity and readability of the manuscript text. After using these tools, we reviewed and edited all content as needed and take full responsibility for the integrity and accuracy of the final manuscript.

## Data sharing and Code Availability

The data used in this study are available to Truveta subscribers through Truveta Studio (studio.truveta.com). To support reproducibility while protecting data privacy, the code supporting the findings of this study will be made publicly available at https://github.com/TruvetaResearchPublic/kccq_score_estimation upon publication, along with synthetic (pseudo) inputs and documentation to reproduce the analyses. Because the shared inputs are synthetic, the reproduced numerical results may differ from those reported here, but the analytical workflow and overall approach will be the same.

## Supplemental Material

Supplemental Methods S1-8

Table 1-2

Figure 1-3

References cited only in the Supplemental Material are included in the main reference list.

## Notes

### Competing Interest Statement

The authors have declared no competing interest.

### Clinical Trial

N/A

### Author Declarations

This study used fully de-identified data and did not involve interaction with human subjects. Therefore, it was determined to be exempt from Institutional Review Board (IRB) review in accordance with applicable regulations.

